# No Increased Risk of Major Adverse Cardiovascular Events following Nicotinamide Exposure

**DOI:** 10.1101/2024.09.16.24313743

**Authors:** Lee Wheless, Ranya Guennoun, Basia Michalski-McNeely, Katlyn M Gonzalez, Rachel Weiss, Siwei Zhang, Lydia Yao, Chris Madden, Hua-Chang Chen, Jefferson L Triozzi, Ran Tao, Otis Wilson, Quinn S Wells, Adriana Hung, Kristin Bibee, Rebecca I Hartman, Yaomin Xu, VA Million Veteran Program

**Affiliations:** Tennessee Valley Healthcare System VA Medical Center, Vanderbilt University Medical Center; Dermatology Departments of Division of Epidemiology; Medicine, Division of Epidemiology; Washington University in St. Louis, Department of Medicine, Division of Dermatology; Vanderbilt University School of Medicine; Vanderbilt University Medical Center Department of Biostatistics; State University of New York Downstate College of Medicine; Vanderbilt University Medical Center Department of Medicine, Division of Nephrology and Hypertension; Vanderbilt University Medical Center Department of Medicine, Division of Cardiovascular Medicine; University of Virginia School of Medicine Department of Dermatology; VA Boston Healthcare System; Brigham and Women’s Hospital Department of Dermatology

**Keywords:** nicotinamide, major adverse cardiovascular events, skin cancer, epidemiology, cohort study

## Abstract

**IMPORTANCE:** Nicotinamide metabolites have recently been implicated in increased risk of major cardiovascular events (MACE). Supportive data about clinical risk of MACE for nicotinamide users is lacking.

**OBJECTIVE:** To determine whether nicotinamide use results in an increase of MACE.

**DESIGN, SETTING, PARTICIPANTS:** Retrospective cohort study of two patient cohorts, Vanderbilt University Medical Center (VUMC) and Military Veteran Program (MVP). The risk of MACE in patients exposed to nicotinamide was compared to the risk of MACE in unexposed patients. In the VUMC cohort, 1228 patients were exposed to nicotinamide based on keyword entry for “nicotinamide” or “niacinamide” and hand-review of charts, while 253 were unexposed but had documented recommendation for use. In the MVP cohort, there were 1594 with exposure to nicotinamide propensity score matched to 2694 without exposure.

**EXPOSURES:** The primary exposure for the VUMC cohort was a confirmed exposure to nicotinamide in chart review. The primary exposure for the MVP cohort was medication entry for “nicotinamide” or “niacinamide”.

**MAIN OUTCOME(S) AND MEASURE(S):** The primary outcome was development of MACE based on a validated phenotype.

**RESULTS:** Between both cohorts, 6039 patients were included, of whom 5125 were male with a mean age of 63.2 years. Neither cohort had significant differences in mean age, sex, race and ethnicity between the nicotinamide exposed and unexposed groups. In the VUMC cohort, there was no significant association between nicotinamide exposure and the primary outcome of MACE (HR 0.76, 95% CI 0.46 – 1.25, p = 0.28). MACE prior to nicotinamide exposure was strongly associated with subsequent MACE (HR 9.01, 95% CI 5.90 – 13.70, p < 0.001). In the MVP cohort, we adjusted for MACE risk factors as potential confounding variables and saw no significant association between nicotinamide exposure and MACE (HR 1.00 95% CI 0.75 – 1.32), while history of prior MACE remained strongly associated with subsequent MACE (HR 9.50, 95% CI 6.38 – 14.1).

**CONCLUSIONS AND RELEVANCE:** In this retrospective cohort study of 6039 adults from two different patient populations, we found no increased risk of MACE in patients with nicotinamide exposure.

## INTRODUCTION

Nicotinamide is a water-soluble, activated form of vitamin B3 that is widely used as a chemopreventive agent against skin cancer^1–4^. More recently, there have been concerns regarding the safety of nicotinamide as it relates to the risk of major adverse cardiovascular events (MACE)^5^. Levels of two nicotinamide metabolites, N1-methyl-2-pyridone-5-carboxamide (2PY) and N1-methyl-4-pyridone-3-carboxamide (4PY), were associated with common variants in the aminocarboxymuconate semialdehyde decarboxylase (*acmsd*) gene, which is responsible for nicotinamide metabolism. The mechanism of this risk modulation is believed to be increased expression of soluble vascular cell adhesion molecule -1 (VCAM-1) due to excess levels of 2PY and 4PY^5^. Increased VCAM-1 has been shown in turn to increase the risk of MACE^6^. The study by Ferrell et al. did not actually examine the real-world experience of patients on nicotinamide, and it focused primarily on niacin, an upstream molecule in nicotinamide metabolism. Whereas nicotinamide is used primarily for chemoprevention of skin cancer or in treatment regimens for bullous pemphigoid, niacin is used to lower cholesterol. As a result, patients with an indication for niacin treatment might have an elevated baseline risk of MACE compared to those exposure to nicotinamide. We conducted this study to assess the risk of developing MACE following exposure to nicotinamide specifically.

## METHODS

### Vanderbilt Cohort

Following approval from the Vanderbilt University Medical Center (VUMC) Institutional Review Board (IRB)(# VR54787), we assembled an electronic health record (EHR) cohort from VUMC’s de-identified EHR research database the Synthetic Derivative (SD) ^7^. The primary exposure was a keyword or medication entry for “nicotinamide” or its other name “niacinamide” (hereafter referred to simply as “nicotinamide”). Patients with oral nicotinamide exposure were confirmed using hand-review of charts. The date of first exposure or first mention of indication was considered baseline. There was insufficient granularity in the data to determine duration of exposure or cumulative dose in the Vanderbilt cohort. The primary outcome was development of MACE, as defined by a modified validated phenotype that included International Classification of Disease (ICD) codes (ICD-9 410-411, ICD-10 I21, I22, I24), Current Procedural Terminology (CPT) codes (3353-6, 33510-4, 33516-19, 33521-3, 92980-6, C1874-7), and laboratory values^8^. Briefly, this required the occurrence of at least two ICD or CPT codes for MACE, plus an elevated troponin level within a five-day window. The original phenotype used CPT codes that have since been retired from use, and both these and the newer codes were included. Elevated troponin was defined as any assay being above its reported upper limit of normal. The date of code entry was used to determine to time of event for each of these outcomes. Censoring occurred at either death or the date of last patient contact as documented by hand-review of the chart.

### Million Veteran Program (MVP) Cohort

This study utilized MVP version 23 release 1 and was approved by the central Veterans Administration IRB (1750541). Nicotinamide exposure was determined by entry in the medication list. Because the MVP cohort had more granular medication data, we included only those Veterans with at least 360 days of medication exposure, equivalent to four 90-day prescriptions of 500mg twice daily. We included only those with at least one year of data prior to first nicotinamide exposure to assess risk of prior MACE. Nicotinamide levels increase rapidly in the blood following oral administration, and trials have shown clinical effects can be seen as early as one month after initiation of treatment^9,10^. Therefore the at-risk window began 30 days after the first exposure.

Unexposed controls were selected from the remaining cohort and matched on propensity score. Propensity score matching is used to reduce selection bias by creating sets balanced on factors associated with the treatment indication^11^. The model was of the form: exposed ∼ year of birth + gender + race + ethnicity + minimum year in the EHR + maximum year in the EHR + date of first code for hypertension + date of first code for congestive heart failure + date of first code for diabetes + date of first code for atherosclerotic cardiovascular disease + date of first code for smoking + date of first code for valvular heart disease + history of bullous pemphigoid + date of first code for bullous pemphigoid + total number of skin cancers + age at first skin cancer. A complete list of codes used for MACE risk factors can be found in Supplemental Table 1. Validated phenotypes did not exist for all of these, and the simple presence of two or more ICD codes was used a proxy measure for history of disease. Skin cancers were counted using the validated co-occurrence of both an ICD and CPT code for skin cancer on the same day^12^. Exact matching was required for year of birth, gender, race, ethnicity, maximum year in the EHR, minimum year in the EHR, history of bullous pemphigoid and number of skin cancers by grouping that approximated the overall distribution of skin cancer counts (0, 1-3, 4-9, 10-15, 16-25, 26 or more). Due to the highly skewed range of individual skin cancer counts, exact count matching led to numerous unmatched individuals and was abandoned. Missing dates were coded with an indicator variable outside of the range of the data to remove it from matching closely to a neighbor with an existing date. Propensity score matching was conducted using the MatchIt package in R using 2:1 matching without replacement, using the nearest neighbor method and distance calculated by generalized linear model. A standardize mean difference plot for the matching is shown in Supplemental Figure 1.

Unlike the Vanderbilt cohort, there was no date of indication in the chart for potential unexposed patients. Therefore, this date had to be imputed. Nicotinamide is commonly used either as a treatment for bullous pemphigoid or as prophylaxis against skin cancer development. There is no set number of skin cancers after which patients should be initiated on nicotinamide, and some start is before the first skin cancer, while others do not start it until they had developed multiple already. Patients were rigorously matched on these indications for nicotinamide treatment, in addition to demographics, age, timing of indication in the EHR, and temporal risk factors for MACE, so we used the date of first exposure for each exposed patient for the matched unexposed patients in each stratum.

### Statistical Analyses

Differences between groups were tested using Student’s T test and Chi-squared tests for continuous and categorical variables, respectively. The tidycmprisk package in R was used to construct conditional and nonconditional competing hazards models in the MVP and VUMC cohorts, respectively, to measure the subdistribution hazard of each outcome with the competing hazard of non-MACE death and calculate hazard ratios (HR) and 95% confidence intervals (CI) ^13,14^. Death occurring on the same day as MACE was coded as MACE. Preliminary analyses suggested a strong association between prior and subsequent MACE, so patients with MACE prior to the date of first nicotinamide exposure were classified using a binary indicator variable, and the first MACE following exposure was considered the outcome. Grey’s tests were used to test differences in time to events for patients with exposure to nicotinamide. All analyses were conducted using R v4.1.2 with p < 0.05 considered statistically significant. This manuscript was prepared according to the Strengthening the Reporting of Observational studies in Epidemiology (STROBE) guidelines^15^.

## RESULTS

### Vanderbilt Cohort

In the Vanderbilt cohort, of the 3,874,690 patients in the SD, there were 3,111 with a keyword or prescription for nicotinamide. Upon hand review of the charts, we excluded nearly half of these as they only had mention of nicotinamide in a standardize patient information form, had small amounts of nicotinamide included in their insulin or total parenteral nutrition, or used a topical formulation. We further excluded 113 of these due to having follow-up time <30 days after first exposure. This left 1,228 with exposure to nicotinamide, including 85 with MACE, and 253 with documented indication that was never started, including 19 with MACE. Patients were predominantly non-Hispanic white with roughly equal numbers of males and females in both the nicotinamide exposed and unexposed groups (Table 1). There was no significant difference in age at baseline between the groups (58.4 vs 57.4, p = 0.44). In univariate analyses, there was no significant association between nicotinamide exposure and MACE (HR 0.76, 95% CI 0.46 – 1.25, p = 0.28), however prior MACE was strongly associated with subsequent MACE (HR 9.01, 95% CI 5.90 – 13.70, p < 0.001). In a model containing both variables, the univariate estimates were minimally changed, with nicotinamide still not associated (HR 0.92, 95% CI 0.58 – 1.48, p = 0.74) and prior MACE strongly associated (HR 8.94, 95% CI 5.91 – 13.50, p < 0.001). Further adjusting for age at baseline, race, and ethnicity yielded similar results (nicotinamide HR 0.84, 95% CI 0.53 – 1.33, p = 0.45; prior MACE HR 4.68, 95% CI 2.97 – 7.36, p < 0.001). Based on the cumulative incidence plot (Figure 1), there appeared to be an interaction between nicotinamide exposure and prior MACE. We tested this and did find evidence of a protective effect of nicotinamide among patients with prior MACE (interaction HR 0.20, 95% CI 0.06 – 0.64 p = 0.006).

**Table 1.**
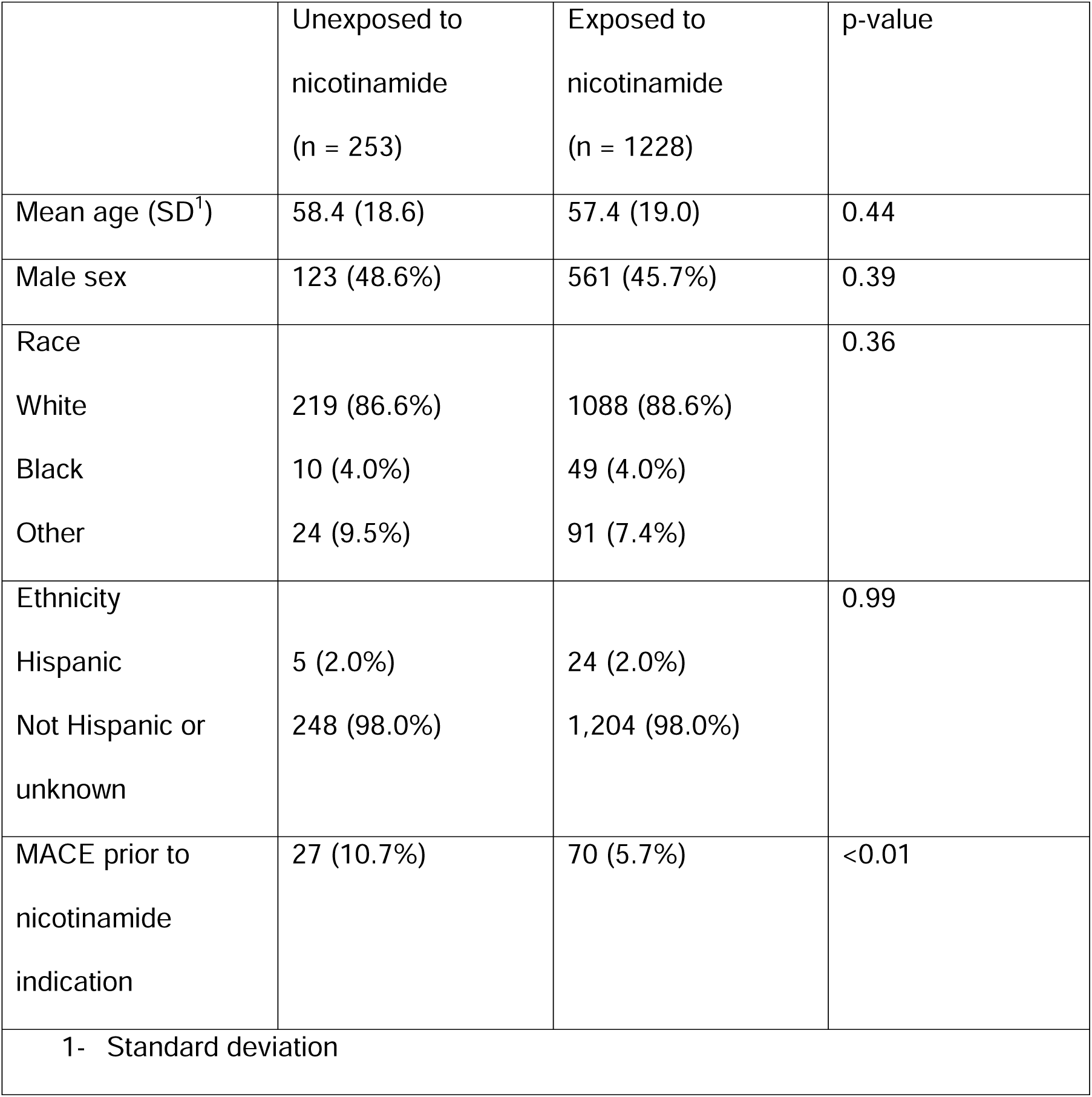
Characteristics of patients in the Vanderbilt cohort included in this study.

**Figure 1.**
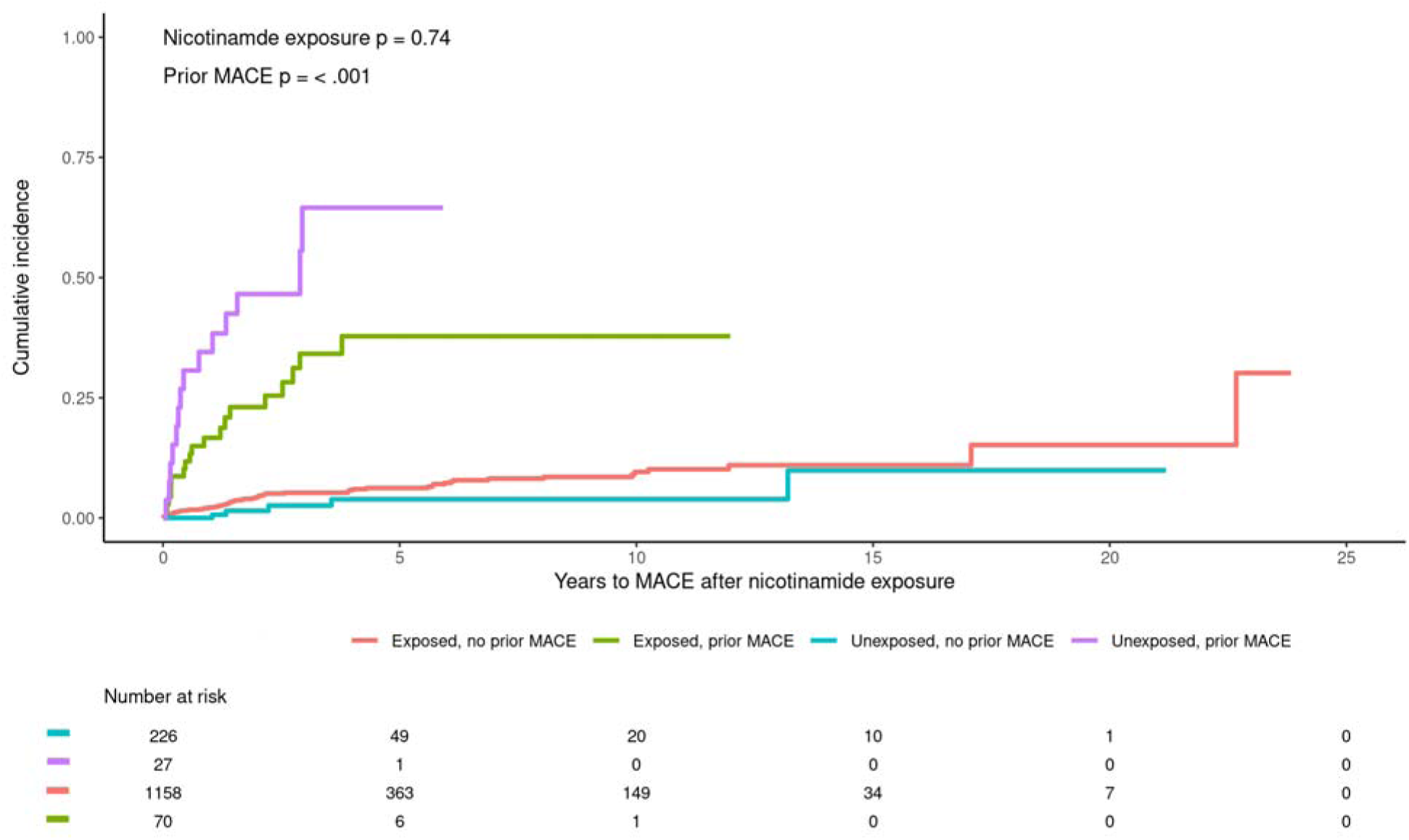
Cumulative incidence of MACE in the Vanderbilt cohort based on nicotinamide exposure and history of prior MACE.

### MVP Cohort

In MVP, of the 963,753 Veterans, there were 4,889 with at least one prescription for nicotinamide. Propensity score matching was performed prior to applying additional exclusion criteria and yielded no significant deviations from balance across any of the variables included in the model (Supplemental Figure 1). The majority of these had the final sample included 1,594 exposed and 2,964 matched unexposed Veterans. There were 224 exposed patients that could be matched to only one unexposed patient. With 1,594 Veterans, 2:1 matching, and assuming a 17-year median MACE-free survival for unexposed Veterans, we estimated that we would have 80% power to detect a hazard ratio of 1.14, which is well below the 1.39 – 2.02 range presented in Ferrell et al.

As expected, the MVP cohort was predominantly male sex (4,441, 97.4%), and white race (4,382, 96.1%)(Table 2). The mean age was 68.5 +/- 8.8 years. A majority had a history of hypertension (74.6%), and a large proportion had atherosclerotic cardiovascular disease (35.7%) or diabetes (43.7%). There was a very strong association between history of MACE and subsequent MACE (HR 38.7, 95% CI 31.3 – 47.8, p < 0.001). Similar to the VUMC cohort, in the cumulative incidence plot there appeared to be a greater reduction in risk among those with prior MACE, so we modeled an interaction between exposure and prior MACE but this was not significant (Fine test p = 0.40)(Figure 2). We further adjusted for any residual confounding based on hypertension (HR 1.30, 95% CI 0.86 – 1.96), congestive heart failure (HR 2.06, 95% CI 1.46 – 2.91), valvular heart disease (HR 1.14, 95% CI 0.82 – 1.58), diabetes (HR 1.01, 95% CI 0.75 – 1.34), atherosclerotic cardiovascular disease (HR 1.95, 95% CI 1.43 – 2.68), or number of skin cancers (HR 0.98, 95% CI 0.97 – 1.00), and exposure to nicotinamide remained non-significant (HR 1.00 95% CI 0.75 – 1.32) while history of prior MACE remained strongly associated with subsequent MACE (HR 9.50, 95% CI 6.38 – 14.1).

**Table 2.** Characteristics of patients in the MVP cohort included in this study.

|  | Unexposed to<br>nicotinamide<br>(n = 2964) | Exposed to<br>nicotinamide<br>(n = 1594) | p-value |
| --- | --- | --- | --- |
| Mean age (SD<sup>1</sup>) | 68.5 (8.8) | 68.4 (8.7) | 0.60 |
| Mean age at first<br>skin cancer (SD<sup>1</sup>) | 65.1 (7.7) | 65.3 (8.0) | 0.08 |
| Male sex | 2887 (97.4%) | 1554 (97.5%) | 0.94 |
| Race |  |  | 0.65 |
| White | 2856 (96.4%) | 1526 (95.7%) |  |
| Black | 70 (2.4%) | 37 (2.3%) |  |
| Other | 38 (1.3%) | 31 (1.9%) |  |
| Ethnicity |  |  | 0.29 |
| Hispanic | 53 (1.8%) | 39 (2.4%) |  |
| Not Hispanic or<br>unknown | 2911 (98.2%) | 1555 (97.6%) |  |
| History of: |  |  |  |
| Hypertension | 2218 (74.8%) | 1184 (74.3%) | 0.71 |
| CHF<sup>2</sup> | 474 (16.0%) | 262 (16.4%) | 0.73 |
| ASCVD<sup>3</sup> | 1051 (35.5%) | 577 (36.2%) | 0.64 |
| DM<sup>4</sup> | 1292 (43.6%) | 699 (43.9%) | 0.89 |
| Valvular disease | 404 (13.6%) | 257 (16.1%) | 0.03 |
| Smoking | 795 (26.8%) | 383 (24.0%) | 0.04 |
| Bullous pemphigoid | 30 (1.0%) | 24 (1.5%) | 0.19 |
| Total number of skin |  |  | 0.04 |
| cancers |  |  |  |
| 0 | 637 (21.5%) | 326 (20.5) |  |
| 1-3 | 768 (25.9%) | 388 (24.3) |  |
| 4-9 | 787 (26.6%) | 404 (25.3) |  |
| 10-15 | 373 (12.6%) | 207 (13.0%) |  |
| 16-25 | 245 (8.3%) | 154 (9.7%) |  |
| 26+ | 154 (5.2%) | 115 (7.2%) |  |
| MACE<sup>5</sup> prior to<br>nicotinamide<br>indication | 72 (2.4%) | 32 (2.0%) | 0.42 |
| 1 – standard deviation<br>2 – congestive heart failure<br>3 – atherosclerotic cardiovascular disease<br>4 – diabetes mellitus<br>5 – major adverse cardiovascular events |  |  |  |

**Figure 2.**
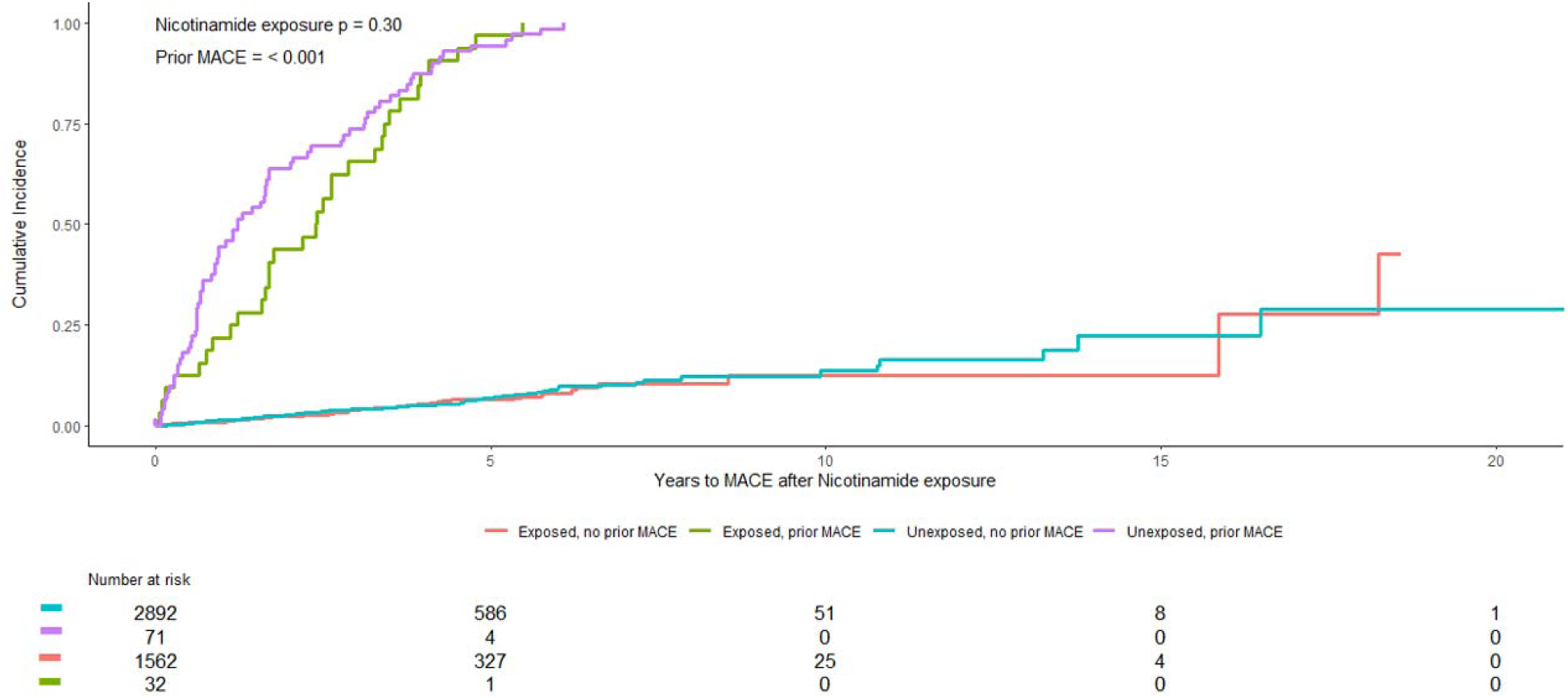
Cumulative incidence of MACE in the MVP cohort based on nicotinamide exposure and history of prior MACE.

Twenty percent of patients had no recorded history of skin cancer treatment. Of these, 48 had a history of bullous pemphigoid, and 915 had no known indication for nicotinamide. Because there was the potential for confounding based on an unmeasured variable, we conducted sensitivity analyses excluding these patients. The results did not change with nicotinamide exposure still showing a non-significant protective effect (HR 0.87, 95% CI 0.68 – 1.10, p = 0.2) and prior MACE showing a strong association (HR 34.5, 95% CI 27.6 – 43.3, p < 0.001).

## DISCUSSION

In this retrospective multi-site EHR cohort study of greater than 2,800 patients with exposure to nicotinamide, we observed no significant association between nicotinamide exposure and risk of MACE. Instead, we saw that patients with an elevated baseline risk with history of prior MACE were likely to develop subsequent MACE. These results were consistent across two different cohorts with different demographic composition and different methods to estimate the date of first exposure or indication for nicotinamide.

Our results are contrary to the conclusions by Ferrell et al that excess metabolites from niacin and, via the same pathway, nicotinamide convey increased risk of cardiovascular disease^5^. In that study, the authors use a proxy measure for both niacin and nicotinamide, which have different indications for use. Whereas niacin can lower cholesterol, nicotinamide does not have this property. As a result, patients treated with niacin likely experience an elevated baseline risk of MACE compared to patients treated with nicotinamide only. Moreover, in the validation cohorts the authors considered quartiles of metabolites and did not measure directly which patients were taking niacin or nicotinamide. In each cohort, only the highest quartile showed any significant increased risk of MACE, showing a lack of a dose-response that would generally be expected for a causal exposure. There is the potential for confounding by indication, as patients with elevated risk of MACE and higher cholesterol might be more likely to be taking niacin and have elevated metabolite levels. In the MVP cohort, we therefore rigorously controlled not only for indication for nicotinamide, but also for risk factors for MACE and their timing to ensure that baseline risks for MACE were equivalent among the exposed and unexposed patients. After this matching, patients with elevated baseline risks experienced MACE at higher rates, regardless of exposure to nicotinamide.

In the VUMC cohort, low numbers prevented us from controlling as aggressively for MACE risk factors. A strength of this cohort, though, was that we had the exact date of indication for patients who were not exposed to nicotinamide and thus did not have to impute it. This cohort was younger on average and had far more females than the MVP cohort. When we stratified by those with prior MACE, we found the exact same results as in the larger MVP cohort with imputed dates, providing strong validation across distinct populations and study designs. We did see a significant reduction in risk of subsequent MACE among patients taking nicotinamide, although there is the potential for confounding by other MACE risk factors that were better accounted for in the MVP cohort that showed a similar but non-significant reduction in risk of subsequent MACE in this group.

The usual indication for nicotinamide is multiple skin cancers. In the MVP cohort, we matched on this variable to control for indication. Nevertheless, many patients in this cohort had no history of skin cancer or other documented indication for use. We conducted sensitivity analyses excluding these patients due to fears of confounding by an unmeasured indication and observed no difference in the overall association.

Our study had several limitations. In the VUMC cohort, there were few patients with documented indication who did not ultimately start nicotinamide. Despite a smaller sample size, we were still able to detect significant risks and interactions within the data. In the MVP cohort, there was no documentation of indication for the unexposed patients, and dates had to be imputed based on complex matching. The cohorts also had different composition, with MVP tending to be older with a larger proportion of males. Despite these differences in study design and cohort composition, the two showed remarkably consistent findings in that patients with prior MACE were much more likely to develop subsequent MACE, and exposure to nicotinamide did not appear to increase risk of MACE, rather it potentially might even have a protective one in patients with prior MACE. While we were sufficiently powered to detect associations of the previously reported magnitude, we did not have adequate power to claim equivalence. However, in the MVP cohort there was a non-significant 11% reduction in risk of MACE with nicotinamide exposure, so increasing our power would be more likely to show a protective effect than a harmful one. There is the potential that unexposed controls were overfit by the propensity score matching, especially seeing the very high HR in the group with prior MACE. First, the risk of subsequent MACE we observed is in line with prior estimates^16^. Second, because the start date had to be imputed following matching, we could not match on prior MACE, but instead matched on multiple risk factors for MACE. There was no difference in the number of individuals with prior MACE among those exposed or unexposed to nicotinamide in the MVP cohort, suggesting that the matching for this very strong risk factor was appropriate. These two points also argue that the unvalidated approach to measuring comorbidities and indications is unlikely to have significantly biased the results, especially considering the high prevalence of MACE risk factors among the Veteran population. The choice of imputing a date of first exposure from the matched exposed patient instead of other methods such as regression could be questioned. Because we incorporated so many time-dependent variables in the propensity score model, the regression models often failed to converge or were too poor-performing for use. Instead, the fact that our findings from the MVP cohort were closely replicated in the Vanderbilt cohort suggests this was a reasonable approach.

## CONCLUSIONS

In this study of two cohorts with different demographic compositions, different baseline risks for MACE, and different analytic approaches, we observed no increased risk of MACE following exposure to nicotinamide. Rather, patients with prior MACE and elevated baseline risk for MACE were more likely to experience subsequent MACE. Our study was underpowered to conclude clinically-meaningful equivalence between exposed and unexposed groups, but our data should reassure clinicians that nicotinamide does not appear to convey increased risks of MACE.

## Supporting information

for the VA Million Veteran Program

## Abbreviations

2PY: N1-methyl-2-pyridone-5-carboxamide
4PY: N1-methyl-4-pyridone-3-carboxamide
*acmsd*: aminocarboxymuconate semialdehyde decarboxylase
CI: confidence interval
CPT: Current Procedural Terminology
EHR: electronic health record
HR: hazard ratio
ICD: International Classification of Disease
IRB: Institutional Review Board
MACE: major adverse cardiovascular events
MVP: Million Veteran Program
SD: Synthetic Derivative
STROBE: Strengthening the Reporting of Observational studies in Epidemiology (STROBE)
VCAM-1: vascular cell adhesion molecule -1
VUMC: Vanderbilt University Medical Center

## Funding and Acknowledgements

*This research is based on data from the Million Veteran Program, Office of Research and Development, Veterans Health Administration, and was supported by awards IK2 CX002452 (Wheless) and CX002531 (Hartman). Please see Supplementary File 1 for the full MVP Core Acknowledgements. Dr. Hartman is supported by the Department of Defense under award number W81XWH2110819. The project described was supported by CTSA award No. **UL1 TR002243** from the National Center for Advancing Translational Sciences. Its contents are solely the responsibility of the authors and do not necessarily represent official views of the National Center for Advancing Translational Sciences, the National Institutes of Health, the Department of Veterans Affairs or the United States Government*.

## Data sharing statement

Individual-level data from both cohorts are access-restricted and cannot be shared

## KEY POINTS

### Question

Does nicotinamide exposure increase risk of major adverse cardiovascular events (MACE)?

### Findings

In two cohorts, nicotinamide exposure was not associated with increased risk of MACE. Rather, patients with prior MACE were likely to develop subsequent MACE.

### Meaning

In a real-world sample, nicotinamide does not appear to convey increased risk of MACE

