## Supplemental Figure 1 for "No Increased Risk of Major Adverse Cardiovascular Events following Nicotinamide Exposure"

Supplemental figure 1. Standardized Mean Difference plot showing the mean differences between exposed patients and the pool of unexposed patients in variables before (white circles) and after (black circles) propensity score matching. The dotted vertical line is 0.05 and the vertical solid line is at 0.1

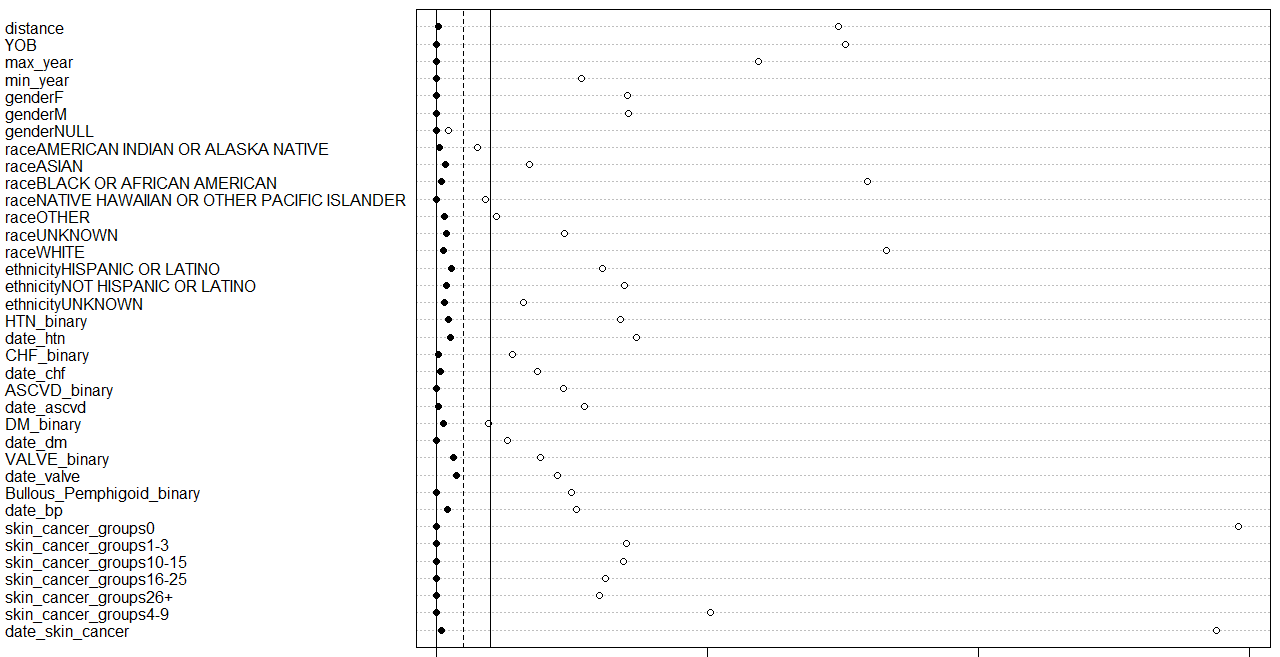
