## Supplemental Table 1 for "No Increased Risk of Major Adverse Cardiovascular Events following Nicotinamide Exposure"

Supplemental Table 1. ICD codes used to identify MACE comorbidities and nicotinamide indications.

| Disease | Codes |
| --- | --- |
| Hypertension | ICD9: 401, 401.1, 401.9, 402, 402.01, 402.1, 402.11, 402.9, 402.91, 403, 403.01, 403.1, 403.11, 403.9, 403.91, 404, 404.01, 404.02, 404.03, 404.1, 404.11, 404.12, 404.13, 404.9, 404.91, 404.92, 404.93, 405.01, 405.09, 405.11, 405.19, 405.91, 405.99, 416  ICD10: I11.0, I11.9, I12.0, I12.9, I13.0, I13.10, I13.11, I13.2, I15.0, I15.1, I15.2, I15.8, I15.9, I16.0, I16.1, I16.9, I1A.0, I27.0, I27.20, I27.21, I27.22, I27.23, I27.24, I27.29 |
| Chronic kidney disease | ICD9: 249.4, 250.4, 250.41, 250.42, 250.43, 403, 403.01, 403.1, 403.11, 403.9, 403.91, 404, 404.01, 404.02, 404.03, 404.1, 404.11, 404.12,  404.13, 404.9, 404.91, 404.92, 404.93, 585.1, 585.2, 585.3, 585.4, 585.5, 585.6, 585.9  ICD10: E08.22, E09.22, E10.22, E11.22, E13.22, I12.0, I12.9, I13.0, I13.10, I13.11, I13.2, N18.2, N18.30, N18.31, N18.32, N18.4, N18.5, N18.6, N18.9, N18.1 |
| Diabetes mellitus | ICD9: 249, 249.01, 249.1, 249.11, 249.2, 249.21, 249.3, 249.31, 249.4, 249.41, 249.5, 249.51, 249.6, 249.61, 249.7, 249.71, 249.8, 249.81, 249.9, 249.91, 250, 250.01, 250.02, 250.11, 250.12, 250.13, 250.2, 250.21, 250.22, 250.23, 250.3, 250.31, 250.32, 250.33, 250.4, 250.41, 250.42, 250.43, 250.5, 250.51, 250.52, 250.53, 250.6, 250.61, 250.62, 250.71, 250.72, 250.73, 250.8, 250.81, 250.82, 250.83, 250.9, 250.91, 250.92, 250.93, 362.01, 362.02, 362.03, 362.04, 362.05, 362.06, 362.07  ICD10: E08.3551, 250.03, 250.10, 250.63, 250.70, E08.01, E08.10, E08.11, E08.21, 08.22, E08.29, E08.311, E08.319, E08.3211, E08.3212, E08.3213, E08.3219, E08.3291, E08.3292, E08.3293, E08.3299, E08.3311, E08.3312, E08.3313, E08.3319, E08.3391, E08.3392, E08.3393, E08.3399, E08.3411, E08.3412, E08.3413, E08.3419, E08.3491, E08.3492, E08.3493, E08.3499, E08.3511, E08.3512, E08.3513, E08.3519, E08.3521, E08.3522, E08.3523, E08.3529, E08.3531, E08.3532, E08.3533, E08.3539, E08.3541, E08.3542, E08.3543, E08.3549, E08.3552, E08.3553, E08.3559, E08.3591, E08.3592,  E08.3593, E08.3599, E08.36, E08.37X1, E08.37X2, E08.37X3, E08.37X9, E08.39, E08.40, E08.41, E08.42, E08.43, E08.44, E08.49, E08.51, E08.52, E08.59, E08.610, E08.618, E08.620, E08.621, E08.622, E08.628, E08.630, E08.638, E08.641, E08.649, E08.65, E08.69, E08.8, E08.9, E09.00, E09.01, E09.10, E09.11, E09.21, E09.22, E09.29, E09.311, E09.319, E09.3211, E09.3212, E09.3213, E09.3219, E09.3291, E09.3292, E09.3293, E09.3299, E09.3311, E09.3312, E09.3313, E09.3319, E09.3391, E09.3392, E09.3393, E09.3399, E09.3411, E09.3412, E09.3413, E09.3419, E09.3491, E09.3492, E09.3493, E09.3499, E09.3511, E09.3512, E09.3513, E09.3519, E09.3521, E09.3522, E09.3523, E09.3529, E09.3531, E09.3532, E09.3533, E09.3539, E09.3541, E09.3542, E09.3543, E09.3549, E09.3551, E09.3552, E09.3553, E09.3559, E09.3591, E09.3592, E09.3593, E09.3599, E09.36, E09.37X1, E09.37X2, E09.37X3, E09.37X9, E09.39, E09.40, E09.41,  E09.42, E09.43, E09.44, E09.49, E09.51, E09.52, E09.59, E09.610, E09.618, E09.620, E09.621, E09.622, E09.628, E09.630, E09.638, E09.641, E09.649, E09.65, E09.69, E09.8, E09.9, E10.10, E10.11, E10.21, E10.22, E10.29, E10.311, E10.319, E10.3211, E10.3212, E10.3213, E10.3219, E10.3291, E10.3292, E10.3293, E10.3299, E10.3311, E10.3312, E10.3313, E10.3319, E10.3391, E10.3392, E10.3393, E10.3399, E10.3411, E10.3412, E10.3413, E10.3419, E10.3491, E10.3492, E10.3493, E10.3499, E10.3511, E10.3512, E10.3513, E10.3519, E10.3521, E10.3522, E10.3523, E10.3529, E10.3531, E10.3532, E10.3533, E10.3539, E10.3541, E10.3542, E10.3543, E10.3549, E10.3551, E10.3552, E10.3553, E10.3559, E10.3591, E10.3592, E10.3593, E10.3599, E10.36, E10.37X1, E10.37X2, E10.37X3, E10.37X9, E10.39, E10.40, E10.41, E10.42, E10.43, E10.44, E10.49, E10.51, E10.52, E10.59, E10.610, E10.618, E10.620, E10.621, E10.622, E10.628, E10.641, E10.649, E10.65, E10.69, E10.8, E10.9, E10-630, E10-638, E11.00, E11.01, E11.10, E11.11, E11.21, E11.22, E11.29, E11.311, E11.319, E11.3211, E11.3212, E11.3213, E11.3219, E11.3291, E11.3292, E11.3293, E11.3299, E11.3311, E11.3312, E11.3313, E11.3319, E11.3391, E11.3392, E11.3393, E11.3399, E11.3411, E11.3412, E11.3413, E11.3419, E11.3491, E11.3492, E11.3493, E11.3499, E11.3511, E11.3512, E11.3513, E11.3519, E11.3521, E11.3522, E11.3523, E11.3529, E11.3531, E11.3532, E11.3533, E11.3539, E11.3541, E11.3542, E11.3543, E11.3549, E11.3551, E11.3552, E11.3553, E11.3559, E11.3591, E11.3592, E11.3593, E11.3599, E11.36, E11.37X1, E11.37X2, E11.37X3, E11.37X9, E11.39, E11.40, E11.41, E11.42, E11.43, E11.44, E11.49, E11.51, E11.52, E11.59, E11.610, E11.618, E11.620, E11.621, E11.622, E11.628, E11.641, E11.649, E11.65, E11.69, E11.8, E11.9, E11-630, E11-638, E13.00, E13.01, E13.10, E13.11, E13.21, E13.22, E13.29, E13.311, E13.319, E13.3211, E13.3212, E13.3213, E13.3219, E13.3291, E13.3292, E13.3293, E13.3299, E13.3311, E13.3312, E13.3313, E13.3319, E13.3391, E13.3392, E13.3393, E13.3399, E13.3411, E13.3412, E13.3413, E13.3419, E13.3491, E13.3492, E13.3493, E13.3499, E13.3511, E13.3512, E13.3513, E13.3519, E13.3521, E13.3522, E13.3523, E13.3529, E13.3531, E13.3532, E13.3533, E13.3539, E13.3541, E13.3542, E13.3543, E13.3549, E13.3551, E13.3552, E13.3553, E13.3559, E13.3591, E13.3592, E13.3593, E13.3599, E13.36, E13.37X1, E13.37X2, E13.37X3, E13.37X9, E13.39, E13.40, E13.41, E13.42, E13.43, E13.44, E13.49, E13.51, E13.52, E13.59, E13.610, E13.618, E13.620, E13.621, E13.622, E13.628, E13.641, E13.649, E13.65, E13.69, E13.8, E13.9, E13-630, E13-638, E08.00 |
| Smoking | ICD9: 305.1, 305.10, 305.11, 305.12, 305.13  ICD10: F17.200, F17.201, F17.203, F17.208, F17.209, F17.210, F17.211, F17.213, F17.218, F17.219, F17.290, F17.291, F17.293, F17.298, F17.299, T65.21, T65.22, T65.29, Z72.0, Z71.6, Z87.8 |
| Atherosclerotic cardiovascular disease | ICD9: 414, 414.01, 414.02, 414.03, 414.04, 414.05, 414.06, 414.07, 414.2, 414.3, 414.4  ICD10: I25.10, I25.110, I25.112, I25.118, I25.119, I25.700, I25.701, I25.702, I25.708, I25.709, I25.710, I25.711, I25.712, I25.718, I25.719, I25.720, I25.721, I25.722, I25.728, I25.729, I25.730, I25.731, I25.732, I25.738, I25.739, I25.750, I25.751, I25.752, I25.758, I25.759, I25.760, I25.761, I25.762, I25.768, I25.769, I25.790, I25.791, I25.792, I25.798, I25.799, I25.810, I25.811, I25.812, I25.82, I25.83, I25.84 |
| Congestive heart Failure | ICD9: 398.91, 402.01, 402.11, 402.91, 414, 414.01, 414.02, 414.03, 414.04, 414.05, 414.06, 414.07, 414.2, 414.3, 414.4, 428, 428.1, 428.2, 428.21, 428.22, 428.23, 428.3, 428.31, 428.32, 428.33, 428.4, 428.41, 428.42,  428.43, 428.9  ICD10: I11.0, I50.1, I50.20, I50.21, I50.22, I50.23, I50.30, I50.31, I50.32, I50.33, I50.40, I50.41, I50.42, I50.43, I50.9, I09.81 |
| Valvular heart disease | ICD9: 36.42, 98.84, 112.81, 115.04, 115.14, 115.94, 394, 394.1, 394.2, 394.9, 395, 395.1, 395.2, 395.9, 396, 396.1, 396.2, 396.3, 396.8, 396.9, 397, 397.1, 397.9, 421, 421.1, 421.9, 424, 424.1, 424.2, 424.3, 424.9, 424.91, 424.99, 746, 746.01, 746.02, 746.09, 746.1, 746.2, 746.3, 746.4, 746.5, 746.6, 746.7, 746.81, 746.83  ICD10: A32.82, A39.51, A52.03, A54.83, B33.21, B37.6, I01.1, I05.0, I05.1, I05.2, I05.8, I05.9, I06.0, I06.1, I06.2, I06.8, I06.9, I07.0, I07.1, I07.2, I07.8, I07.9, I08.0, I08.1, I08.2, I08.3, I08.8, I08.9, I09.0, I09.1, I09.2, I09.89, I09.9, I33.0, I33.9, I34.0, I34.1, I34.2, I34.81, I34.89, I34.9, I35.0, I35.1, I35.2, I35.8, I35.9, I36.0, I36.1, I36.2, I36.8, I36.9, I37.0, I37.1, I37.2, I37.8, I37.9, I38, I39, M32.11, Q22.0, Q22.1, Q22.2, Q22.3, Q22.4, Q22.5, Q22.6, Q22.8, Q22.9, Q23.0, Q23.1, Q23.2, Q23.3, Q23.4, Q23.8, Q23.9, Q24.3, Q24.4 |
| Bullous pemphigoid | ICD9: 694, 694.5  ICD10: L12.0 |
