## Supplementary material for "No Increased Risk of Major Adverse Cardiovascular Events following Nicotinamide Exposure": for the VA Million Veteran Program

**Core Acknowledgements for Publications**

**MVP Program Office**

- Sumitra Muralidhar, Ph.D., Program Director

US Department of Veterans Affairs, 810 Vermont Avenue NW, Washington, DC 20420

- Jennifer Moser, Ph.D., Associate Director, Scientific Programs

US Department of Veterans Affairs, 810 Vermont Avenue NW, Washington, DC 20420

- Jennifer E. Deen, B.S., Associate Director, Cohort & Public Relations

US Department of Veterans Affairs, 810 Vermont Avenue NW, Washington, DC 20420

**MVP Executive Committee**

-	Co-Chair: Philip S. Tsao, Ph.D.

VA Palo Alto Health Care System, 3801 Miranda Avenue, Palo Alto, CA 94304

-	Co-Chair: Sumitra Muralidhar, Ph.D.

US Department of Veterans Affairs, 810 Vermont Avenue NW, Washington, DC 20420

-	J. Michael Gaziano, M.D., M.P.H.

VA Boston Healthcare System, 150 S. Huntington Avenue, Boston, MA 02130

-	Elizabeth Hauser, Ph.D.

Durham VA Medical Center, 508 Fulton Street, Durham, NC 27705

-	Amy Kilbourne, Ph.D., M.P.H.

VA HSR&D, 2215 Fuller Road, Ann Arbor, MI 48105

-	Michael Matheny, M.D., M.S., M.P.H.

VA Tennessee Valley Healthcare System, 1310 24th Ave. South, Nashville, TN 37212

-	Dave Oslin, M.D.

Philadelphia VA Medical Center, 3900 Woodland Avenue, Philadelphia, PA 19104

-	Deepak Voora, MD

Durham VA Medical Center, 508 Fulton Street, Durham, NC 27705

**MVP Co-Principal Investigators**

- J. Michael Gaziano, M.D., M.P.H.

VA Boston Healthcare System, 150 S. Huntington Avenue, Boston, MA 02130

- Philip S. Tsao, Ph.D.

VA Palo Alto Health Care System, 3801 Miranda Avenue, Palo Alto, CA 94304

**MVP Core Operations**

- Jessica V. Brewer, M.P.H., Director, MVP Cohort Operations

VA Boston Healthcare System, 150 S. Huntington Avenue, Boston, MA 02130

- Mary T. Brophy M.D., M.P.H., Director, VA Central Biorepository

VA Boston Healthcare System, 150 S. Huntington Avenue, Boston, MA 02130

- Kelly Cho, M.P.H, Ph.D., Director, MVP Phenomics

VA Boston Healthcare System, 150 S. Huntington Avenue, Boston, MA 02130

- Lori Churby, B.S., Director, MVP Regulatory Affairs

VA Palo Alto Health Care System, 3801 Miranda Avenue, Palo Alto, CA 94304

- Scott L. DuVall, Ph.D., Director, VA Informatics and Computing Infrastructure (VINCI)

VA Salt Lake City Health Care System, 500 Foothill Drive, Salt Lake City, UT 84148

- Saiju Pyarajan Ph.D., Director, Data and Computational Sciences

VA Boston Healthcare System, 150 S. Huntington Avenue, Boston, MA 02130

- Robert Ringer, Pharm.D., Director, VA Albuquerque Central Biorepository

New Mexico VA Health Care System, 1501 San Pedro Drive SE, Albuquerque, NM 87108

- Luis E. Selva, Ph.D., Director, MVP Biorepository Coordination

VA Boston Healthcare System, 150 S. Huntington Avenue, Boston, MA 02130

- Shahpoor (Alex) Shayan, M.S., Director, MVP PRE Informatics

VA Boston Healthcare System, 150 S. Huntington Avenue, Boston, MA 02130

- Brady Stephens, M.S., Principal Investigator, MVP Information Center

Canandaigua VA Medical Center, 400 Fort Hill Avenue, Canandaigua, NY 14424

- Stacey B. Whitbourne, Ph.D., Director, MVP Cohort Development and Management

VA Boston Healthcare System, 150 S. Huntington Avenue, Boston, MA 02130

**MVP Publications and Presentations Committee**

- Co-Chair: Themistocles L. Assimes, M.D., Ph. D

VA Palo Alto Health Care System, 3801 Miranda Avenue, Palo Alto, CA 94304

- Co-Chair: Adriana Hung, M.D.; M.P.H

VA Tennessee Valley Healthcare System, 1310 24^th^ Ave. South, Nashville, TN 37212

- Co-Chair: Henry Kranzler, M.D.

Philadelphia VA Medical Center, 3900 Woodland Avenue, Philadelphia, PA 19104
